# Subcordal Stenosis: A Glucocorticoid-Responsive Subtype of Autoimmune Laryngeal Stenosis

**DOI:** 10.1101/2025.07.25.25332220

**Authors:** Brendan Denvir, Bridget Burgess, Kevin Motz, Simon RA Best, Lee M Akst, Brendan Antiochos, Philip Seo, Alexander T Hillel

## Abstract

**Objective:** As our understanding of autoimmune laryngeal stenosis evolves, distinguishing patients who may benefit from systemic immunosuppression versus those needing only local treatment is increasingly important. In this study, we identify a distinct subset of autoimmune laryngeal stenosis characterized by edema of the inferior true vocal folds that extends to the superior aspect of the cricoid cartilage, termed “subcordal stenosis.” The objective of this study is to characterize the clinical presentation and treatment outcomes of subcordal stenosis and compare it to typical autoimmune-related subglottic stenosis (AI-SGS).

**Methods:** We conducted a retrospective review of patients with laryngeal stenosis evaluated by both rheumatology and otolaryngology at our institution to identify two groups: patients with subcordal stenosis and those with typical AI-SGS. Data on immunosuppressive treatments and airway dilation procedures were collected. Time to first dilation was compared between groups.

**Results:** Among 49 patients with laryngeal stenosis, 11 had subcordal involvement. Five of these also had subglottic disease, while six had isolated subcordal stenosis. Kaplan-Meier analysis showed significantly longer time to first dilation in patients with subcordal involvement (median 792 vs. 44 days; p = 0.048). They also underwent fewer dilations within two years (median 0 vs. 1; p = 0.05).

**Conclusion:** Among laryngeal stenosis patients referred to rheumatology, those with subcordal involvement experienced fewer dilations and longer intervals before first dilation compared to those with typical AI-SGS. These findings suggest that subcordal stenosis may represent a distinct, glucocorticoid-responsive phenotype within autoimmune laryngeal stenosis, with implications for treatment selection and multidisciplinary care.

## Introduction

Subglottic stenosis (SGS) is characterized by narrowing of the airway at the level of the cricoid cartilage and proximal trachea.^1^ Acquired SGS can result from a variety of causes, including iatrogenic injury, infections, systemic autoimmune disease, and idiopathic etiologies.^1,2^ In the absence of a clear infectious or iatrogenic cause, the differential diagnosis commonly narrows to idiopathic SGS (iSGS) and autoimmune SGS (AI-SGS). Distinguishing between these two entities is critical, as it directly informs clinical management.

iSGS accounts for approximately 20% of SGS cases and occurs in an estimated 1 in 400,000 individuals. It primarily affects Caucasian women between the third and fifth decades of life.^1^ In contrast, the most common autoimmune cause of SGS is granulomatosis with polyangiitis (GPA),^3^ an antineutrophil cytoplasmic antibody (ANCA)-associated vasculitis that classically involves the upper respiratory tract, lungs, and kidneys. GPA can affect all levels of the airway, with tracheal involvement reported in approximately 12–27% of patients.^4–7^ Notably, airway disease may occur even in the absence of severe pulmonary or renal involvement,^4,5,8^ and in some cases, may be the sole manifestation of GPA.^9^

Diagnosing GPA in patients with SGS can be challenging, particularly when extra-otolaryngologic features are absent. ANCA serologies can assist in distinguishing AI-SGS from iSGS, as ANCA positivity supports a diagnosis of GPA.^3^ However, ANCA testing lacks complete specificity and can be positive in several non-vasculitic conditions,^10^ and up to 25% of patients with subglottic stenosis associated with GPA (SGS-GPA) are ANCA negative.^4–8^ Histopathology can help differentiate iSGS from SGS-GPA, but its diagnostic yield is extremely low. Biopsies from patients with tracheal involvement of GPA often show nonspecific inflammation, with diagnostic features such as granulomas, vasculitis, or necrosis being rare.^7,11,12^

Clinical management differs substantially between iSGS and SGS-GPA. In both conditions, serial endoscopic dilations and local corticosteroid injections are commonly employed, while cricotracheal resection or reconstruction may be considered for severe or refractory disease.^4,13–16^ However, SGS-GPA benefits from systemic immunosuppression in addition to procedural treatment. Several retrospective studies have demonstrated that immunosuppression reduces the procedural burden in SGS-GPA.^8,12,17–19^ In contrast, immunosuppressive therapy has been trialed in iSGS with unclear benefit.^20^ Thus, distinguishing between idiopathic and autoimmune SGS is central to determining the appropriate therapeutic approach, particularly in assessing whether immunosuppression is warranted.

In this study, we describe a distinct pattern of laryngotracheal involvement—termed subcordal stenosis—to aid in this diagnostic distinction. This phenotype is characterized by swelling and stenosis originating at the inferior margin of the true vocal folds (TVFs), bounded by the conus elasticus. At times it presents along with the more typical subglottic stenosis that extends into the proximal trachea. In our clinical experience, subcordal stenosis is associated with autoimmune rather than idiopathic laryngeal stenosis and may be uniquely responsive to glucocorticoids compared to typical SGS.

The objective of this study is to characterize subcordal stenosis as a distinct clinical phenotype within autoimmune laryngeal stenosis and to evaluate differences in clinical features and glucocorticoid responsiveness between patients with subcordal stenosis and typical SGS. We conducted a retrospective analysis of patients with suspected autoimmune laryngeal stenosis seen at our medical center, focusing on a cohort of patients that were referred to rheumatology. At our center, patients are referred from otolaryngology to rheumatology when clinical suspicion for autoimmune laryngeal stenosis exists, based on atypical demographics for iSGS, positive ANCA serologies, extra-otolaryngologic manifestations, or inflammatory appearance of the airway. The majority of these patients receive systemic treatment with prednisone, with or without steroid-sparing immunosuppression. In this context, we compared outcomes of patients with subcordal stenosis to those with typical SGS. We hypothesized that patients with subcordal stenosis would be a similar demographic cohort to AI-SGS, have a greater responsiveness to glucocorticoid therapy, and thereby have a lower procedural burden compared with AI-SGS.

## Materials and Methods

### Patient Selection

IRB approval was obtained from the Johns Hopkins University Institutional Review Board prior to beginning this study. Informed consent was waived due to the retrospective nature of the study and minimal risk associated with data collection. Medical records were queried for all patients with ICD-10 codes associated with laryngeal stenosis (J38.6, J39.8, J98.09) who were evaluated by a rheumatologist between January 1, 2013 and December 2024 at Johns Hopkins Medical Institutions. Chart review was performed to confirm a diagnosis of laryngeal stenosis by an otolaryngologist. Patients were excluded if any of the following applied: (1) they were not seen by both the Rheumatology and Otolaryngology departments at Johns Hopkins, (2) there was no chart evidence of laryngeal stenosis, (3) they underwent dilation for laryngeal stenosis prior to presentation at Johns Hopkins, or (4) they had less than six months of follow-up after SGS diagnosis. We excluded patients with prior dilations to standardize the time zero for time-to-event analyses.

Patients with subcordal stenosis were identified through a keyword search of laryngoscopy reports for the terms “subcordal” or “infracordal.” All flagged cases were reviewed by an otolaryngologist to confirm subcordal involvement. Subcordal stenosis was defined as airway narrowing at the level of the inferior TVFs, below the superior margin of the TVFs, and including—but not limited to—the space above the level of the cricoid cartilage (Figure 1).

**Figure 1.**
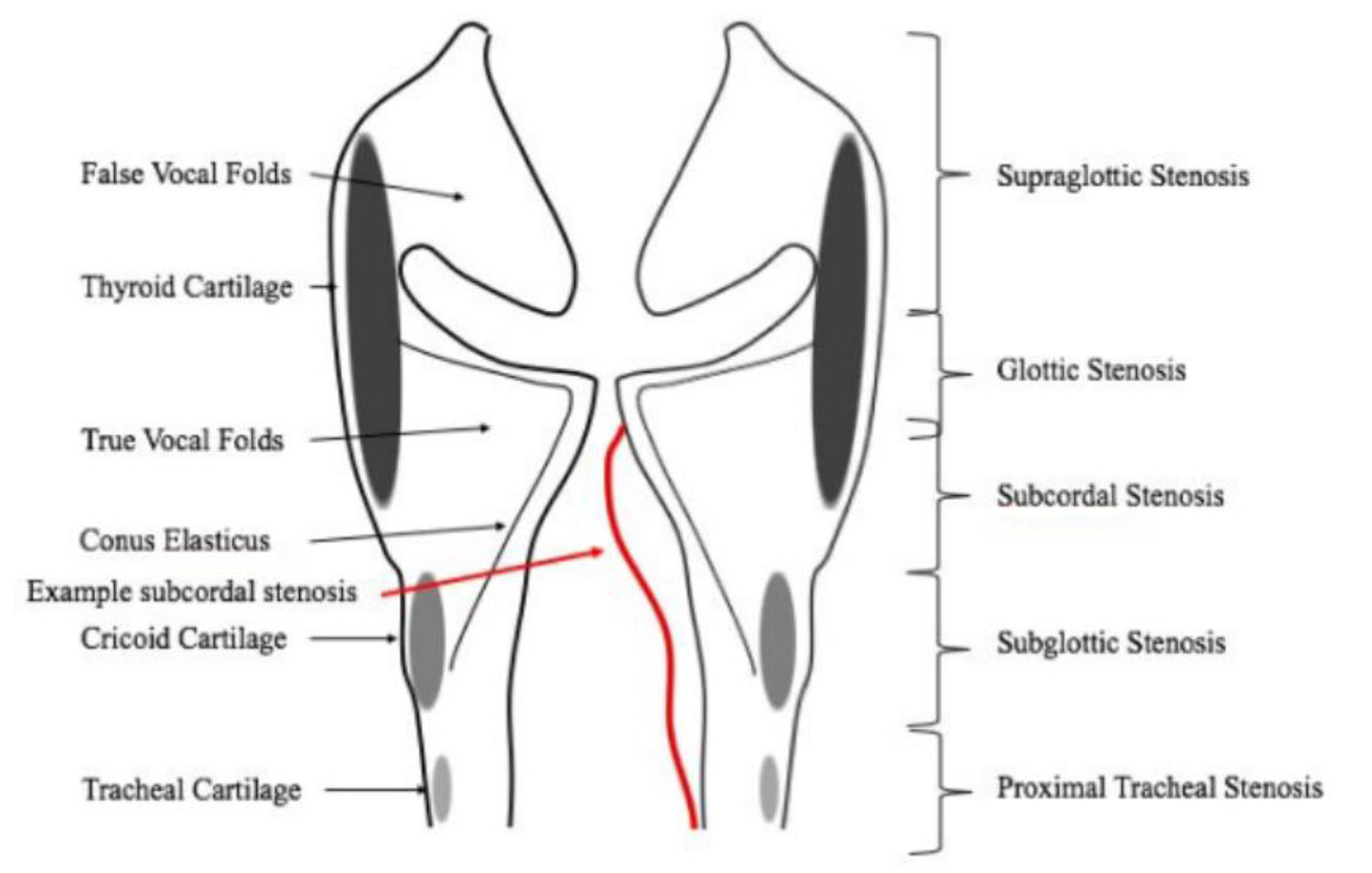
Image of the larynx showing anatomic locations of laryngotracheal stenosis. Subcordal stenosis is highlighted in red beginning at the level of the thyroid cartilage just inferior to the superior aspect of the true vocal folds contained by the conus elasticus.

### Data Collection

We used a standardized data extraction form in REDCap to record data for all patients who met inclusion and exclusion criteria. We collected baseline data on the demographic characteristics and clinical characteristics of the cohort. Demographic data included age, sex, and race. Clinical characteristics included the date of laryngeal stenosis diagnosis, presence of airway symptoms (dyspnea, stridor, dysphonia, hemoptysis), anatomic location of airway stenoses, and descriptors of gross appearance on endoscopy. For patients with a diagnosis of GPA, we collected additional disease-specific variables, including the date of GPA diagnosis, ANCA serologies (ANCA, anti-MPO, and anti-PR3 antibodies), and organ system involvement at diagnosis. Organ manifestations were coded using a structured list derived from the Birmingham Vasculitis Activity Score (BVAS) and the Vasculitis Damage Index (VDI).

Immunosuppressant medication data were abstracted by reviewing rheumatology and otolaryngology notes. We recorded whether patients were treated with prednisone at the time of laryngeal stenosis diagnosis as a binary variable; however, dose and duration were not recorded. We also documented start and stop dates of steroid-sparing immunosuppressants including rituximab, cyclophosphamide, methotrexate, azathioprine, leflunomide, and mycophenolate mofetil.

Procedural interventions were identified through review of clinical and procedural notes. For each patient, we recorded the dates and types of airway procedures, including balloon dilation, laryngotracheoplasty, laryngotracheal resection, and tracheostomy. Adjunctive procedures performed at the time of dilation—such as local steroid injection or laser excision— were also recorded. Standalone steroid injections were not included.

The follow-up period began at the time of laryngeal stenosis diagnosis. The primary outcome was time to first endoscopic dilation. Secondary outcomes included the number of dilations performed within two years of diagnosis, and percentage of patients with zero dilations.

### Data Analysis

All analyses were conducted using R. Descriptive statistics were used to compare baseline characteristics between patients with and without subcordal stenosis. Categorical variables were compared using Fisher’s exact test, and continuous variables using t-tests. Time-to-event analysis was performed using Kaplan–Meier curves, with comparison of median time to first dilation between groups using the log-rank test. The number of dilations within two years was compared using the Wilcoxon rank-sum test, due to the non-parametric distribution of the outcome.

## Results

A total of 49 patients with laryngeal stenosis were included in the study (Figure 2). Of these, 11 patients (22%) had subcordal stenosis, while 38 patients (78%) had typical SGS. Among those with subcordal disease, 5 patients (45%) also had concurrent subglottic involvement, while 6 patients (55%) had isolated subcordal stenosis.

**Figure 2.**
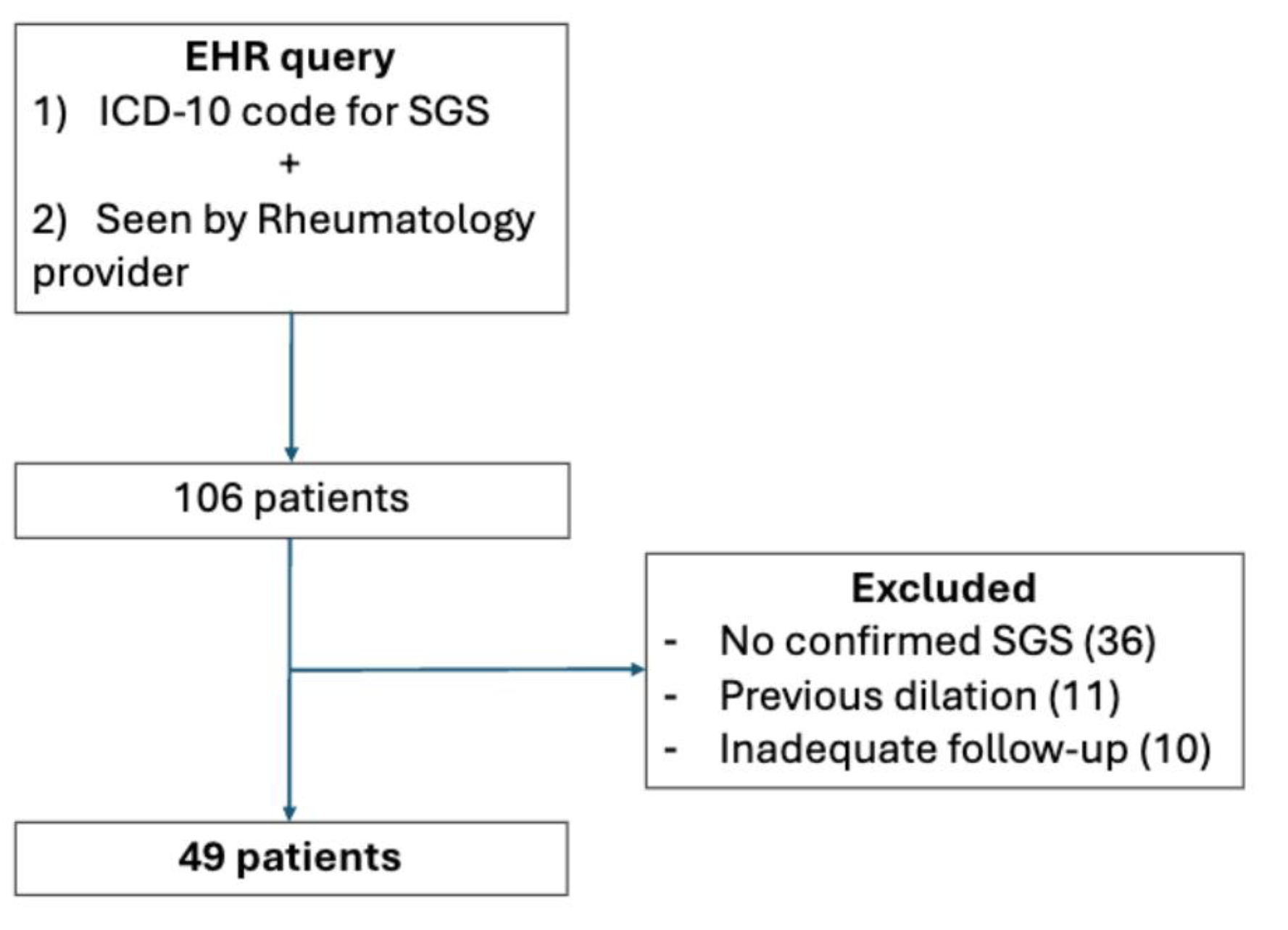
Patient selection flow diagram. We queried medical records for patients with ICD-10 codes for subglottic stenosis (J38.6, J39.8, J98.09) seen by a rheumatologist at Johns Hopkins between January 2013 and December 2024. Otolaryngologist-confirmed SGS was required. Patients with prior dilations were excluded to standardize time zero for time-to-event analyses. Patients with less than 6 months of follow-up were also excluded.

A summary of demographic, clinical, and treatment characteristics is provided in Table 1. GPA was diagnosed by a rheumatologist in 8 of 11 patients (73%) in the subcordal group and in 33 of 38 patients (87%) in the SGS group. ANCA positivity was observed in 67% of the overall cohort, with similar proportions across both groups. Among patients with GPA, there were no differences between groups in the presence of severe GPA disease manifestations.

**Table 1.** Baseline demographic, clinical, and treatment characteristics, and outcome comparisons between laryngeal stenosis patients with and without subcordal involvement.

|  | Subcordal<br>Involvement<br>n=11 | No Subcordal<br>Involvement<br>n=38 | p-value* |
| --- | --- | --- | --- |
| **Demographics** |  |  |  |
| Age, mean (sd) | 44.5 (12.3) | 42.6 (15.3) | 0.69 |
| Female, n (%) | 7 (63.6) | 28 (73.7) | 0.71 |
| Male, n (%) | 4 (36.3) | 10 (26.3) |  |
| **Diagnosis** |  |  |  |
| GPA, n (%) | 8 (72.7) | 33 (86.8) | 0.50 |
| Severe † | 3 (27.3) | 4 (10.5) | 0.18 |
| Non-severe | 5 (45.5) | 29 (76.3) |  |
| ANCA positive | 7 (63.6) | 26 (68.4) | 0.81 |
| Idiopathic SGS, n (%) | 3 (27.3) | 5 (13.2) |  |
| **Airway Involvement** |  |  |  |
| Subglottic | 5 (45.4) | 38 (100) | - |
| Subcordal | 11 (100) | 0 (0) | - |
| **Airway Symptoms, n (%)** |  |  |  |
| Dyspnea | 9 (81.8) | 29 (76.3) | 1 |
| Stridor | 5 (45.5) | 15 (39.5) | 0.74 |
| Cough | 2 (18.2) | 6 (15.8) | 1 |
| Dysphonia | 5 (45.5) | 12 (31.6) | 0.48 |
| **Immunosuppression ‡, n (%)** |  |  |  |
| Prednisone | 10 (90.9) | 27 (71.1) | 0.41 |
| Rituximab | 3 (27.3) | 13 (34.2) | 1 |
| Methotrexate | 2 (18.2) | 13 (34.2) | 0.46 |
| Azathioprine | 3 (27.3) | 4 (10.5) | 0.12 |
| Leflunomide | 3 (27.2) | 3 (7.9) | 0.18 |
| Cyclophosphamide | 1 (9.1) | 1 (2.6) | 0.40 |
| Mycophenolate mofetil | 0 (0) | 1 (2.6) | 1 |

\*Statistical comparisons between groups were made using fisher's exact tests, t-tests and wilcoxon rank sum tests where appropriate.
Immunosuppression indicates medication patients were on in the first year after airway disease diagnosis
† Severe disease defined by the presence of life or organ-threatening manifestations consistent with American College of Rheumatology/Vasculitis Foundation guidelines
‡ Medications received at any point during the follow-up period

Dyspnea was the most common presenting symptom across both groups. Dysphonia was reported in 45% of patients with subcordal stenosis compared to 31% of those with SGS, though this difference was not statistically significant. At the time of airway stenosis diagnosis, 91% of patients with subcordal disease were treated with prednisone, compared to 71% of patients in the SGS group.

Patients with subcordal disease experienced significantly longer time to first dilation compared to those without subcordal involvement. The median time to first dilation was 792 days in the subcordal group versus 44 days in the SGS group (p = 0.048), as shown in Kaplan– Meier curves (Figure 3). Within two years of laryngeal stenosis diagnosis, the median number of dilations was 0 in the subcordal group and 1 in the non-subcordal group (p = 0.05; Table 2). 54.5% of patients in the subcordal group underwent no dilations during this period, compared to 28.9% in the SGS group. No patients in either group underwent tracheostomy, laryngotracheoplasty, or tracheal resection within two-year follow-up period.

**Table 2.**
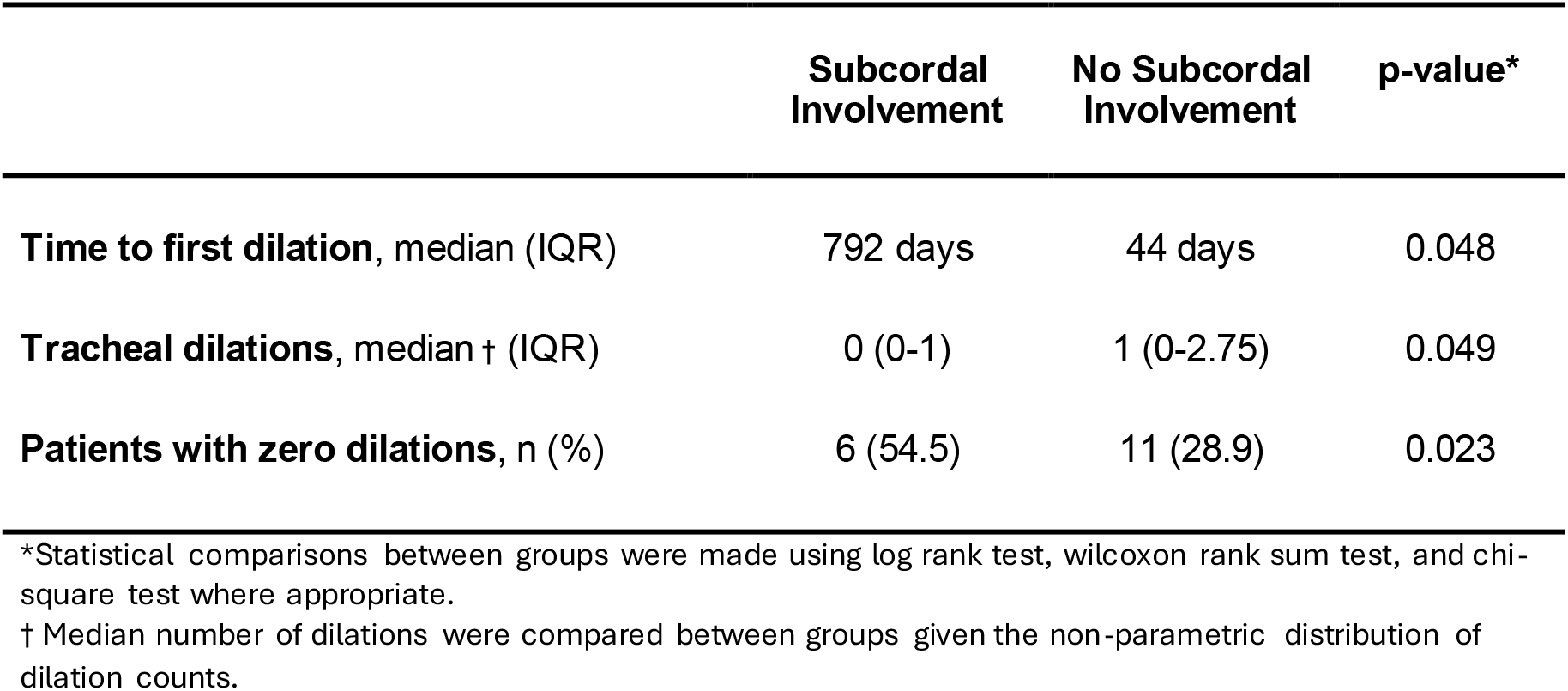
Comparison of procedural outcomes between patients with and without subcordal involvement.

|  | **Subcordal**<br>**Involvement** | **No Subcordal**<br>**Involvement** | **p-value\*** |
| --- | --- | --- | --- |
| **Time to first dilation**, median (IQR) | 792 days | 44 days | 0.048 |
| **Tracheal dilations**, median † (IQR) | 0 (0-1) | 1 (0-2.75) | 0.049 |
| **Patients with zero dilations**, n (%) | 6 (54.5) | 11 (28.9) | 0.023 |

\*Statistical comparisons between groups were made using log rank test, wilcoxon rank sum test, and chi-square test where appropriate.
† Median number of dilations were compared between groups given the non-parametric distribution of dilation counts.

**Figure 3.**
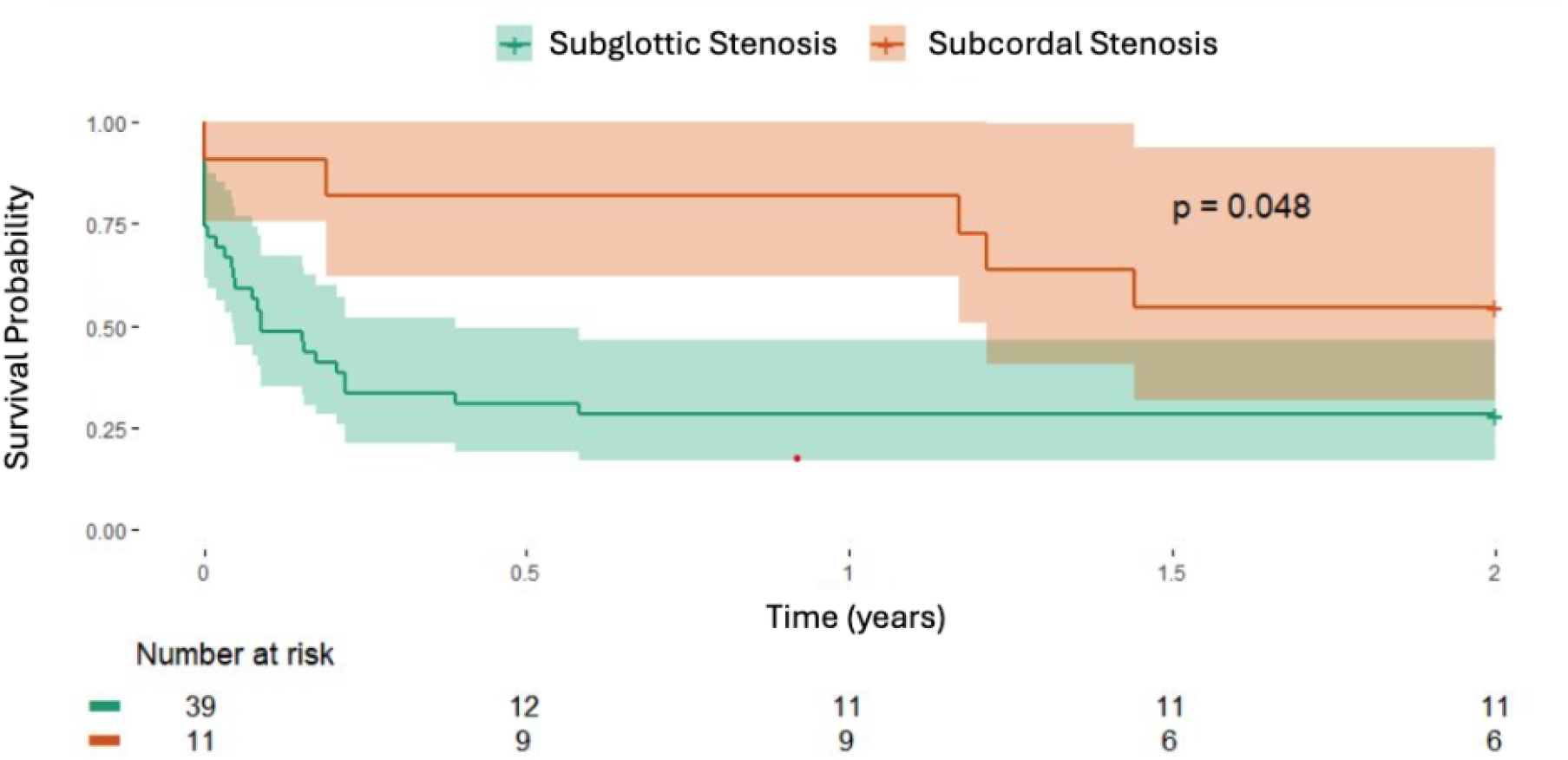
Kaplan-Meier curves for time to first tracheal dilation in patients with subcordal stenosis versus typical subglottic stenosis. Time zero was the date of SGS diagnosis. Patients without dilation were censored at last follow-up or at two years. Comparison by log-rank test; p = 0.048.

## Discussion

As our understanding of autoimmune SGS evolves, it remains critical to distinguish which patients will benefit from systemic immunosuppression versus those who require only local treatment. In this context, we introduce a distinct phenotype of autoimmune laryngeal stenosis, termed subcordal stenosis, characterized by its unique clinical appearance and, based on our experience, increased responsiveness to glucocorticoid therapy. Subcordal stenosis differs from typical SGS both in location and endoscopic findings. Rather than the classic fibrotic stricture of iSGS, subcordal stenosis presents as edema involving the inferior aspect of the true vocal folds, at times with a connection along the posterior laryngeal wall (Figure 4, images d-f). In some cases, this subcordal involvement coexists with more typical SGS. In this study, we evaluated clinical outcomes in 11 patients with subcordal stenosis and compared them to a matched group of patients with subglottic stenosis who had similar demographics, clincal and serologic profiles, and immunosuppressant exposure (Table 1). Patients with subcordal stenosis experienced a longer time to first dilation following diagnosis, underwent fewer dilations within two years, and a greater proportion of these patients required no dilations at all.

**Figure 4.**
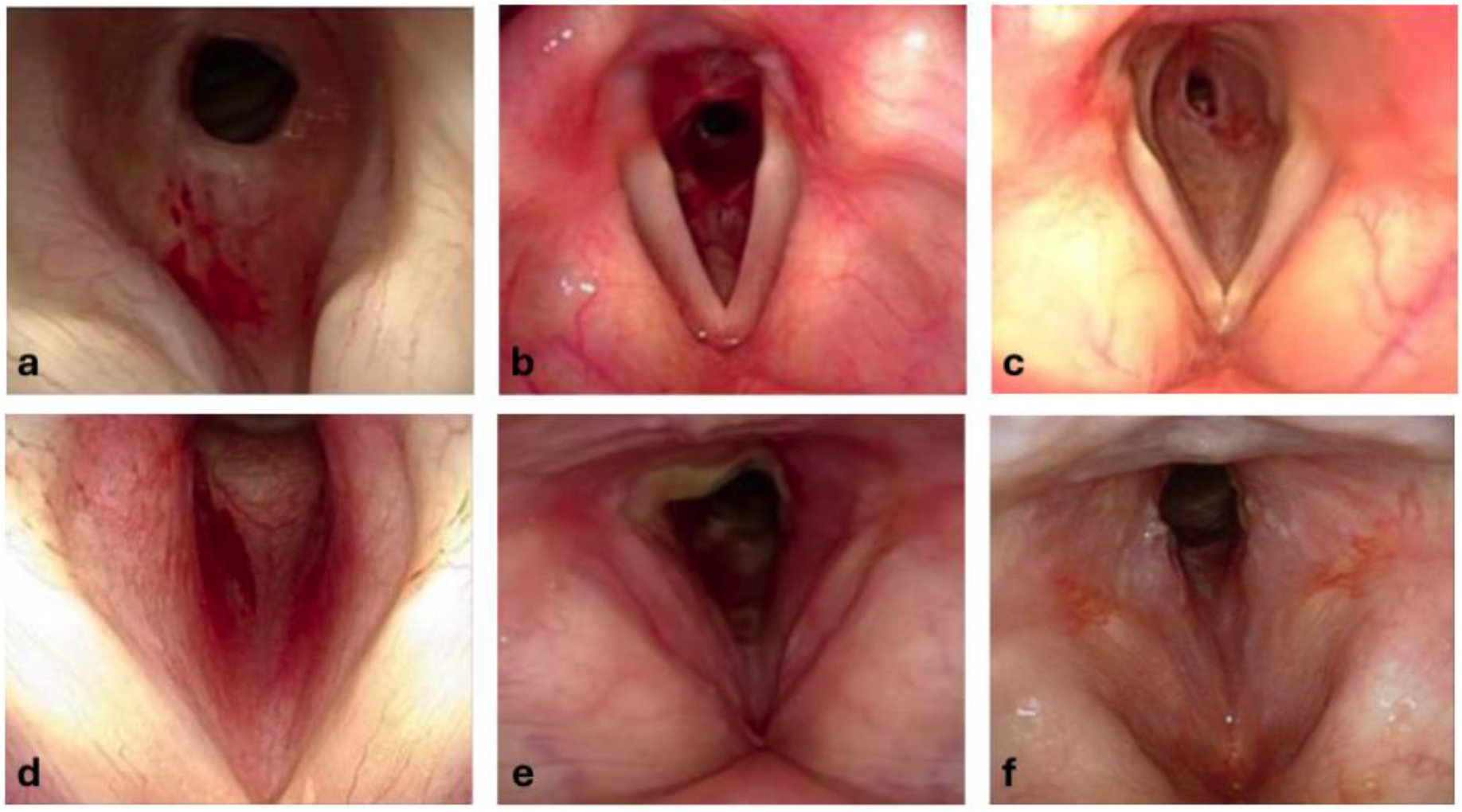
Representative laryngoscopic images of subglottic stenosis versus subcordal stenosis. **Panel A:** Laryngoscopy near view of subglottic stenosis with its typical appearance, starting inferior to the inferior aspect of the true vocal folds, with a fibrotic stricture circumferentially around the airway. **Panel B:** Laryngoscopy far view of GPA-SGS with stenosis starting below the TVFs again with the typical stricture scar appearance. **Panel C:** Laryngoscopy far view of iSGS with 80% stenosis distal to the inferior TVF. Note the similar appearance of subglottic stenosis to that of GPA-SGS. **Panel D:** Laryngoscopy near view of subcordal stenosis showing the fullness and erythema of the inferior aspect of the true vocal folds. It begins inferior to the medial vibratory edge of the TVFs. **Panel E:** Laryngoscopy far view subcordal stenosis starting immediately inferior to the medial edge of the TVFs. Additionally note the exudate in the inter-arytenoid area **Panel F:** Laryngoscopy far view subcordal stenosis with swelling and erythema running into the inferior aspect of the TVF coming up from the subglottic area.

We demonstrate increased systemic immunosuppressive responsiveness in subcordal stenosis relative to typical GPA-SGS. Figure 5 illustrates a representative case demonstrating clinical improvement with glucocorticoid therapy, allowing for symptom control without immediate procedural intervention. This observation raises important mechanistic and clinical questions. It is possible that subcordal stenosis represents an early or more inflammatory stage of autoimmune SGS, offering a therapeutic window during which systemic immunosuppression may reverse airway narrowing and avert procedural intervention. While our cohort primarily included patients with suspected GPA, the potential relevance of subcordal stenosis as a manifestation of other autoimmune or inflammatory airway diseases remains to be explored. It is conceivable that subcordal stenosis reflects a shared airway inflammatory pathway across autoimmune disorders, which warrants further investigation. Other studies have identified distinct features of AI-SGS. For example, Dion et al. reported that posterior glottic involvement is more common in autoimmune SGS than in iSGS.^21^ However, this finding may reflect sequelae of procedural interventions rather than inherent differences in patterns of inflammation. Future studies leveraging airway tissue analysis, including histopathology, may clarify whether subcordal stenosis reflects an early inflammatory phenotype or a distinct immunopathologic process within the spectrum of autoimmune SGS.

**Figure 5.**
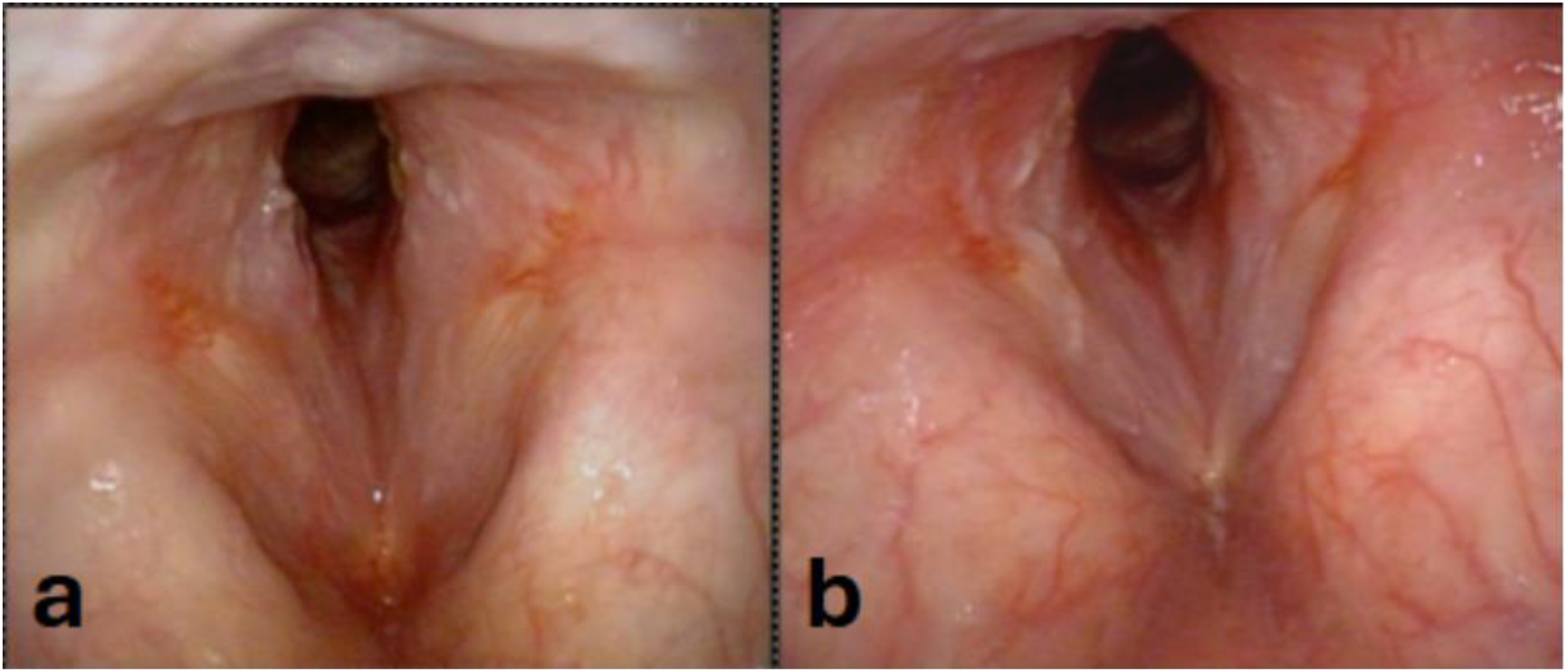
Laryngoscopic images from a patient with subcordal stenosis before and after glucocorticoid treatment. **Panel A**: Subcordal stenosis at presentation, prior to prednisone initiation, showing significant airway narrowing. **Panel B**: Same patient after prednisone treatment, demonstrating continued subcordal stenosis but significantly increased glottic airway.

Although not formally tested in this study, we hypothesize that subcordal stenosis is uniquely associated with underlying autoimmune disease. In our cohort, 73% of patients with subcordal stenosis had a confirmed autoimmune diagnosis—a higher proportion than typically observed in patients with classic SGS, where distinguishing GPA-SGS from iSGS based on laryngoscopic appearance alone can be challenging. Several lines of evidence support this association. First, steroid responsiveness is uncommon in iSGS, which is characterized by fibrotic, non-inflammatory lesions. Second, the demographic profile of subcordal stenosis in our cohort—a 3:4 male-to-female ratio—resembles that of GPA and contrasts sharply with iSGS, which predominantly affects women.

Several limitations should be considered when interpreting the findings of our study. First, there is potential for confounding by treatment practice patterns. At our institution, patients with subcordal stenosis may have undergone fewer procedures due to greater clinical confidence in glucocorticoid responsiveness, which could contribute to the observed differences in dilation rates. Additionally, while we recorded whether patients were treated with glucocorticoids, we did not capture precise data on dose, duration, or adherence, limiting our ability to evaluate the relationship between treatment intensity and clinical outcomes. Future investigations should evaluate whether treatment with steroid-sparing systemic immunosuppression alters disease progression in patients with subcordal disease, reducing the need for procedural intervention and/or glucocorticoid burden. Second, the retrospective design inherently limits control for unmeasured confounding factors, including provider decision-making and disease severity at presentation. Finally, our relatively small sample size and reduced statistical power precluded the use of multivariable modeling or advanced causal inference techniques. Future studies with larger cohorts and prospective data collection will be essential to validate these findings and better define the role of systemic immunosuppression in patients with subcordal stenosis.

Effective management of subcordal stenosis requires close, ongoing collaboration between otolaryngology and rheumatology. Rheumatologists play a critical role in evaluating for systemic autoimmune disease, initiating and monitoring immunosuppressive therapy, and assessing for extra-airway manifestations that may guide treatment intensity. Otolaryngologists are essential for diagnostic recognition through laryngoscopy, longitudinal airway surveillance, and procedural decision-making when interventions are required. Given the potential for subcordal stenosis to signal underlying autoimmune pathology—even in the absence of classic systemic features—early multidisciplinary involvement is essential to ensure timely diagnosis and to optimize outcomes.

## Conclusion

Subcordal stenosis is a rare variant of laryngeal stenosis characterized by edema and narrowing of the inferior aspect of the true vocal folds, often extending to or below the level of the cricoid cartilage and sometimes coexisting with subglottic stenosis. In this study of patients with autoimmune-associated laryngeal stenosis, those with subcordal involvement experienced a significantly longer time to first dilation compared to those without. Our findings suggest that subcordal stenosis represents an inflammatory, autoimmune-driven phenotype that is particularly responsive to systemic glucocorticoid therapy. Unlike GPA-associated subglottic stenosis, which often requires repeated procedural interventions, subcordal stenosis may be more effectively managed with systemic immunosuppression alone, potentially reducing the need for local procedures.

## Data Availability

All data produced in the present study are available upon reasonable request to the authors

